# The Cognitive Connectome in Dementia with Lewy Bodies

**DOI:** 10.1101/2023.11.14.23298515

**Authors:** Roraima Yánez-Pérez, Eloy García-Cabello, Annegret Habich, Nira Cedres, Patricia Diaz-Galvan, Carla Abdelnour, Jon B. Toledo, José Barroso, Daniel Ferreira

**Affiliations:** Division of Clinical Geriatrics, Centre for Alzheimer Research, Department of Neurobiology, Care Sciences, and Society, Karolinska Institutet, Stockholm, Sweden; Department of Clinical Psychology, Psychobiology and Methodology, Faculty of Psychology, University of La Laguna, Canary Islands, Spain; Facultad de Ciencias de la Salud, Universidad Fernando Pessoa Canarias, Las Palmas de Gran Canaria, España; Department of Psychology, Sensory Cognitive Interaction Laboratory (SCI-lab), Stockholm University, Stockholm, Sweden; Department of Radiology, Mayo Clinic, Rochester, MN 55905, United States; Department of Neurology and Neurological Sciences, Stanford University School of Medicine, Stanford, California, USA; Nantz National Alzheimer Center, Stanley H. Appel Department of Neurology, Houston Methodist Hospital, Houston, TX, United States

## Abstract

**Objectives:** Cognition plays a central role for diagnosing and characterizing dementia with Lewy bodies (DLB). However, the complex associations among cognitive functions are largely unknown in DLB. To fill this gap, we compared the cognitive connectome of DLB patients, healthy controls (HC) and patients with Alzheimer’s disease (AD).

**Methods:** We obtained data from the National Alzheimer’s Coordinating Center (NIA/NIH Grant U24-AG072122). We built separate cognitive connectomes for DLB (n=104), HC (n= 3703), and AD (n=1985) groups using pairwise correlations between 24 cognitive variables mapping multiple cognitive functions. The cognitive connectomes in DLB, HC, and AD groups were compared using standard global and nodal graph measures of centrality, integration, and segregation.

**Results:** In global connectome measures, DLB patients showed a higher global efficiency (integration) and lower transitivity (segregation) than HCs and AD. Nodal connectome measures showed a higher global efficiency in most cognitive functions in DLB compared to HCs. Additionally, we found a lower local efficiency (segregation) and nodal strength (centrality) in memory variables and a higher participation coefficient in executive variables (centrality) in DLB compared with both HCs and AD.

**Conclusions:** The cognitive connectome in DLB showed a signature dedifferentiation pattern of aberrant correlations. Executive, processing speed and attention functions played a central role in the cognitive connectome of DLB patients. Furthermore, the role of executive and memory functions in the cognitive connectome distinguished DLB and AD patients. These findings may help advance our understanding of the clinical phenotype in DLB, and continue to improve the challenging differential diagnosis between DLB and AD.

## BACKGROUND

Dementia with Lewy bodies (DLB) is one of the most common neurodegenerative dementias (1). The essential criterion for diagnosing DLB is a progressive cognitive decline (2). Furthermore, cognitive decline not only plays an important role in the diagnosis of DLB but also its differential diagnosis with other dementias (3–5). The typical cognitive profile of DLB includes alterations in attention, executive functions, and visual abilities, while other domains, such as memory, can be involved at later stages of the disease (2).

The traditional approach to investigating cognition in DLB focuses on a particular cognitive function and uses univariate statistical analysis to compare the performance with healthy controls or other relevant dementias such as Alzheimer’s disease (AD) or Parkinson’s disease dementia (6–8). Although the univariate approach can provide information on the cognitive profile of DLB patients, it falls short in elucidating the complex associations between cognitive functions. This limitation has enabled increased interest in multivariate approaches to study cognition in DLB and prodromal stages, promoting a more integrated understanding of cognitive functioning (9–12). In this line, it has been suggested that processing speed mediates the performance in working memory and memory in prodromal DLB (12). It has also been suggested that executive deficits may underlie problems in verbal fluency or that deficits in semantic memory and visual functions may influence the deficits observed in naming (13).

The “cognitive connectome” (14) is a new concept and methodology that comprehensively explains the complex organization and relationships between cognitive functions. Using graph theory analysis on cognitive variables provides rich data on the centrality of specific cognitive functions in the connectome and information on the integration and segregation of cognition (15). While the cognitive connectome has been investigated in normal aging and some disorders such as epilepsy, acquired brain injury, vascular encephalopathy, mild cognitive impairment, or Alzheimer’s disease dementia (14–20), no study to date has characterized the cognitive connectome in DLB. The characterization of the cognitive connectome in DLB could have implications for advancing our understanding of its complex phenotype. Moreover, it could contribute to improve the differential diagnosis of DLB by integrating data profiles instead of evaluating each neuropsychological function separately in an “univariate” manner. Such approach will better align with how clinicians interpret cognitive data.

In this study, we introduced graph theory analysis on cognitive variables in DLB to investigate its cognitive connectome. The first objective was to characterize the cognitive connectome in DLB by comparing it with a group of healthy controls (HCs). The second objective was to compare the cognitive connectome of DLB and AD patients, as this is the most common comparison for differential diagnosis of DLB in the clinical setting. We hypothesized prominent alterations of the cognitive connectome in DLB compared to HCs, particularly involving attention, executive, and visual functions. At the same time, the differences with AD patients would be more modest and likely extend to memory domains.

## METHODS

### Participants

We obtained data from the National Alzheimer’s Coordinating Center (NACC) collected across 32 Alzheimer’s Disease Research Centers between March 2015 and May 2021 (21). Patients aged ≥ 45 years and diagnosed with DLB or AD were included (2,22). Clinical severity was assessed with the Clinical Dementia Rating (CDR) scale (23). Core clinical features of DLB were determined by the clinician’s judgment, including fluctuating cognition, visual hallucinations, probable REM sleep behavior disorder (RBD), and parkinsonism. We also included a group of HCs who demonstrated unimpaired cognition on clinical assessment.

All participants were required to have data available on all the cognitive variables selected to build the connectome (see next section). For all the participants, the exclusion criteria were having a clinical history of bipolar disorder, schizophrenia, delusional disorder, craniocerebral trauma, substance abuse, and uncorrected vision or hearing problems. All participants gave written informed consent, and local Institutional Review Boards approved the study.

### Cognitive measures and construction of the cognitive connectome

The neuropsychological protocol of the NACC database is fully described elsewhere (21). We included 25 cognitive variables mapping multiple cognitive functions, as follows: visuoconstructive functions, visual and verbal episodic memory, processing speed, attention, executive functions, language, and orientation.

Before constructing the cognitive connectome, we carefully inspected the distribution and nature of all the 25 cognitive variables. We inverted the scores when necessary, so that higher scores always indicated a better performance and transformed variables with a heavily skewed distribution. Moreover, since age, sex, and education influence cognitive performance, we adjusted cognitive variables for these confounding factors using multiple linear regression or logistic regression (24). After these steps, data was inspected again to ensure that variables in the new dataset were normally distributed. We observed a variable which did not follow a normal distribution. This variable was finally included as an error variable and was especially informative in a pathological sample. Thus, we used Spearman correlation coefficients to define the edges of the cognitive connectome. At that point, we observed that one variable was barely correlated to the rest of the variables (i.e., repetition errors in phonemic fluency) and was thus excluded from the analysis since we aimed for modelling connectomes with highly correlated variables. The 24 cognitive variables were used to construct a cognitive connectome for each group, i.e., DLB, AD, and HCs (Table 1).

**Table 1.** List of cognitive variables included in the cognitive connectomes.

| **Cognitive functions** | **Most prominent cognitive component** | **Cognitive measures** | **Cognitive variables** |
| --- | --- | --- | --- |
| Visuoconstructive functions | Visuoconstructive abilities | Number of correct elements | Benson Complex Figure- Copying (25,26) |
|  | 2-D visuoconstructive abilities | Number of correct elements<br>Score for a totally correct task | MoCA-Visuoconstructive (27) |
| Visual and verbal episodic memory | Encoding, Retrieval and Storage (Visual memory) | Number of correct elements | Benson Complex Figure-Delayed recall (25,26) |
|  |  | Score for a totally correct task | Benson Complex Figure-Recognition (25,26) |
|  | Encoding, Retrieval and Storage (Verbal memory) | Number of correct words | Craft Story 21-Immediate recall (28) |
|  |  |  | Craft Story 21-Delayed recall (28) |
|  |  |  | MoCA-Immediate recall (27) |
|  |  |  | MoCA-Delayed recall (27) |
|  |  |  | MoCA-Category cue (27) |
|  |  |  | MoCA-Recognition (27) |
| Executive functions | Phonemic (letters), semantic (animals) fluency. Executive control, cognitive inhibition | Number of correct words<br>Number of incorrect words | Verbal fluency-Phonemic; adapted from (29)<br>Verbal fluency-Phonemic-intrusions; adapted from (29)<br>Verbal fluency-Semantic; adapted from (29) |
|  | Executive control, cognitive inhibition | Number of commission errors | Trail Making Test (TMT)-commissions (30) |
|  | Mental flexibility/executive control | Score for a totally correct task | MoCA-Trail (27) |
|  | Mental flexibility/Concept formation | Number of correct items | MoCA-Abstraction (27) |
|  | Working memory: amplitude (forward) and manipulation (backward) | Number of correct items | Digit span forward (25)<br>Digit span backward (25) |
| Processing speed and attention | Focusing/visual tracking | Seconds | TMT-Part A (30) |
|  | Maintenance of attention | Score for a totally correct task | MoCA- Calculation (27) |
|  |  | Number of correct items | MoCA- Letter A (27) |
| Language | Lexical access by visual confrontation | Number of correct items | Multilingual Naming Test (MINT) (31,32) |
|  | Repetition | Number of correct items | MoCA-Repetition (27) |
| Orientation | Temporal and spatial orientation | Number of correct items | MoCA-Orientation (27) |

Following two previous studies on cognitive connectomes in acquired brain injury and epilepsy, we used positive and negative correlations (15,20). Additionally, we excluded self-connections from the correlation matrices. Next, correlation matrices were binarized by thresholding coefficients at a range of densities based on the cognitive connectome of the HC group. Preliminary analysis of the cognitive connectome of HCs showed that MoCA variables tended to form a cluster of correlations, increasing the threshold density at which other nodes became connected. We thus built a simplified connectome without MoCA variables to determine the range of densities. At the minimum density (10%), nodes tended to be connected to at least one other node. At the maximum density (30%), the connectome exhibited a random topology with the small-world index approaching 1. We then compared connectome topologies of DLB patients to HC and AD groups through 1000 nonparametric permutations across the range of densities 10% to 30%, in steps of 1%.

### Graph theory measures

To characterize the DLB cognitive connectome, we calculated centrality, integration, and segregation measures at the global and nodal levels. Among the different graph measures available, we mainly used those that have shown to be stable in previous studies (33) and have previously been used to investigate cognitive connectomes (14). We then calculated the global measures of *average strength* (a measure of centrality)*, global efficiency* (a measure of integration), *transitivity* (a measure of segregation), *and local efficiency* (a measure of segregation) (34). We also calculated the nodal measures of *strength* and *participation* (measures of centrality), *global efficiency, and local efficiency* (34). All measures are fully described in Supplementary Table 1. We calculated all measures on binary networks except for global and nodal *strength* that were calculated on the weighted network (before binarization).

### Statistical Analysis

We used ANOVA for between-group comparisons of demographic and clinical variables and ANCOVA with age, sex, and education as covariates for between-group comparisons of standardized cognitive function scores. We adjusted p-values in post-hoc analyses with Hochberg’s correction for multiple comparisons (35).

Statistical significance was set at *p*<0.05. Results from global graph measures were reported across connectome densities from 10 to 30% in steps of 1%. Global measures with significant differences in ≥5 densities were considered significant. For nodal measures, we applied the false discovery rate (FDR) adjustment at p ≤0.05 (two-tailed) (36). Results from nodal graph measures were also considered across all connectome densities but reported only at the median density (20%).

Statistical analyses were performed using R Studio version 0.99.483 with the ULLRToolbox, SPSS version 25.0, and BRAPH software version 1.0.0 (37).

## RESULTS

### Cohort characteristics

Table 2 shows the main demographic and clinical characteristics of the groups. There were no statistically significant differences in age for DLB compared to HCs and AD. However, there were group differences in sex and education. Hence, we controlled for sex and education when investigating cognitive performance across the three groups but also for age, as it is related to cognitive performance. The DLB and AD groups did not differ in the CDR total score. Figure 1 shows differences in cognitive functions across the groups. Compared to HCs, the DLB group performed significantly worse in all cognitive functions. Compared to AD, the DLB group showed a significantly worse performance in visuoconstructive functions, processing speed, and attention, whereas the AD group showed a worse performance in visual and verbal memory, language, and orientation compared to DLB.

**Figure 1.**
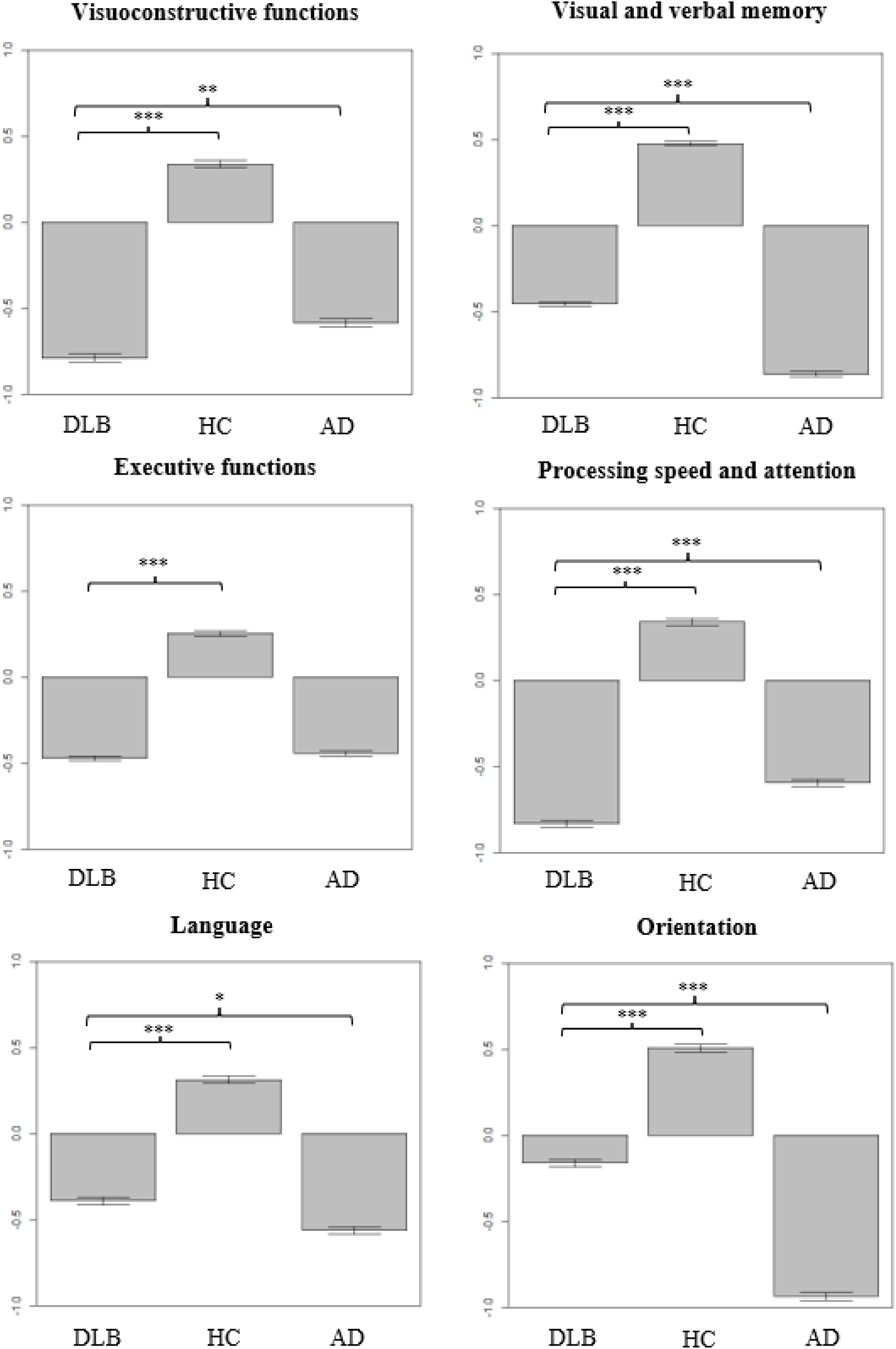
Cognitive performance in the three diagnostic groups. Differences in DLB patients compared to HCs and AD patients in the six cognitive functions were analyzed using ANCOVA with age, sex, and education as covariates. Measures have been adjusted, so a higher score indicates better performance. Whiskers show 95% confidence intervals. *p<0.05; **p<0.01; ***p<0.001.

**Table 2.** Key demographic and clinical characteristics.

|  | DLB |  | HC |  | AD |  | *p*-value |
| --- | --- | --- | --- | --- | --- | --- | --- |
|  | N | Mean<br>(SD)/<br>count<br>(%) | N | Mean<br>(SD)/<br>count<br>(%) | N | Mean<br>(SD)/<br>count<br>(%) |  |
| Age, years<br>min-max | 104 | 71.9<br>(8.5)<br>45-88 | 3703 | 72.2<br>(9.2)<br>45-101 | 1985 | 73.7<br>(9.9)<br>45-103 | N.S. |
| Sex, men (%) | 104 | 89<br>(86%) | 3703 | 1576<br>(43%) | 1985 | 931<br>(47%) | <0.001<br>*ab |
| Education,<br>years | 104 | 16.3<br>(3.1) | 3703 | 16.0<br>(2.9) | 1985 | 15.4<br>(3.0) | <0.05*<br>b |
| CDR-total | 104 | 1 (0.5-1) | 3703 | 0 (0-0) | 1985 | 1 (0.5-1) | <0.001<br>*a |
| Cognitive<br>fluctuations,<br>presence | 102 | 61<br>(60%) | 3702 | 6<br>(0.2%) | 1944 | 78<br>(4%) | <0.001<br>*ab |
| Visual<br>hallucinations,<br>presence | 104 | 45<br>(43%) | 3702 | 8<br>(0.2%) | 1982 | 71<br>(3.6%) | <0.001<br>*ab |
| Probable<br>RBD,<br>presence | 101 | 66<br>(65%) | 3696 | 39<br>(1%) | 1959 | 62<br>(3%) | <0.001<br>*ab |
| Parkinsonian<br>signs,<br>presence | 104 | 85<br>(82%) | 3703 | 127<br>(3%) | 1985 | 210<br>(11%) | <0.001<br>*ab |

Comparisons were established a-priori for DLB vs. HCs and DLB vs. AD. a. Significant differences between DLB and HCs; b. Significant differences between DLB and AD. N.S.: non-significant differences. Abbreviations: CDR, Clinical Dementia Rating scale; RBD, REM sleep behaviour disorders.

### Weighted Correlations Matrices

Figure 2 shows the cognitive connectomes for the three study groups (see Supplementary Figures 1, 2, and 3 for cognitive connectomes with larger size). Visual inspection of the cognitive connectome in DLB showed uniformly weak correlations within and between cognitive functions. Visual inspection of the cognitive connectome in HCs showed the coexistence of both strong and weak correlations within and between cognitive functions. Compared with the HC group, most correlations within and between cognitive functions were generally weaker in DLB. Visual inspection of the cognitive connectome in AD showed a predominance of weak correlations within and between cognitive functions. Comparisons between the DLB and AD groups revealed more moderate differences than the HC group. Most correlations within and between cognitive functions were weaker in DLB compared to AD, especially in visual and verbal memory functions.

**Figure 2.**
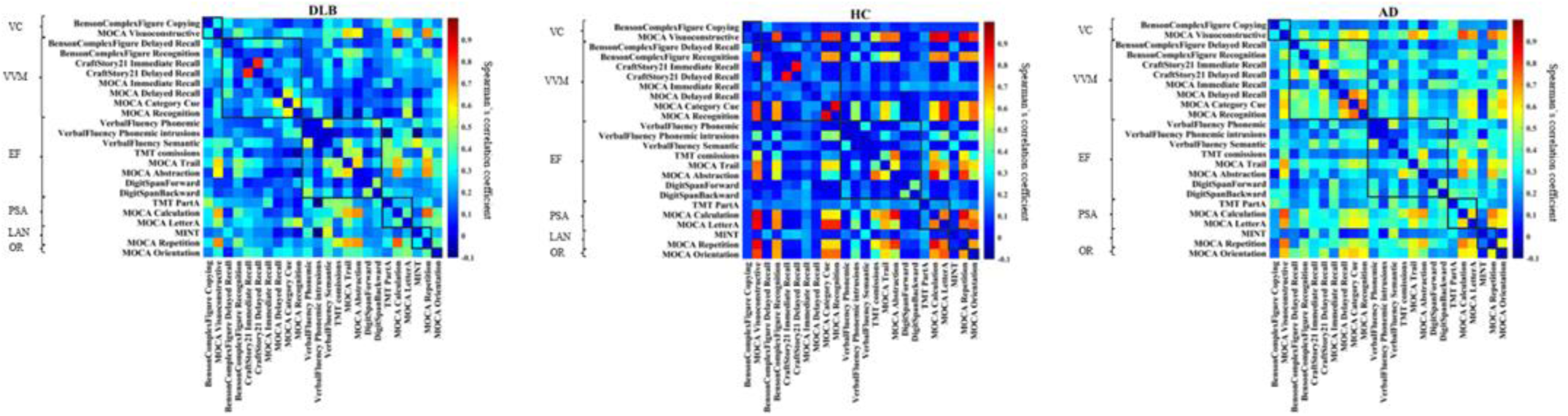
Correlation matrices for each group. Cognitive connectomes for each study group with cognitive variables grouped by cognitive functions. Negative correlations were only observed in DLB and AD groups, but the range of color bar numbers was equalized across all groups to facilitate comparison. VC, visuoconstructive functions; VVM, visual and verbal episodic memory; EF, executive functions; PSA, processing speed and attention; LAN, language; OR, orientation.

### Global connectome measures analysis

Figure 3 shows the quantitative differences of the DLB patients compared to the HC and AD groups in global connectome measures. There were no statistically significant differences in the average strength between the DLB and HC groups. However, DLB patients exhibited a higher global efficiency and a lower local efficiency and transitivity than HCs. Compared with the AD group, DLB patients had a lower average strength, together with a higher global efficiency and a lower transitivity.

**Figure 3.**
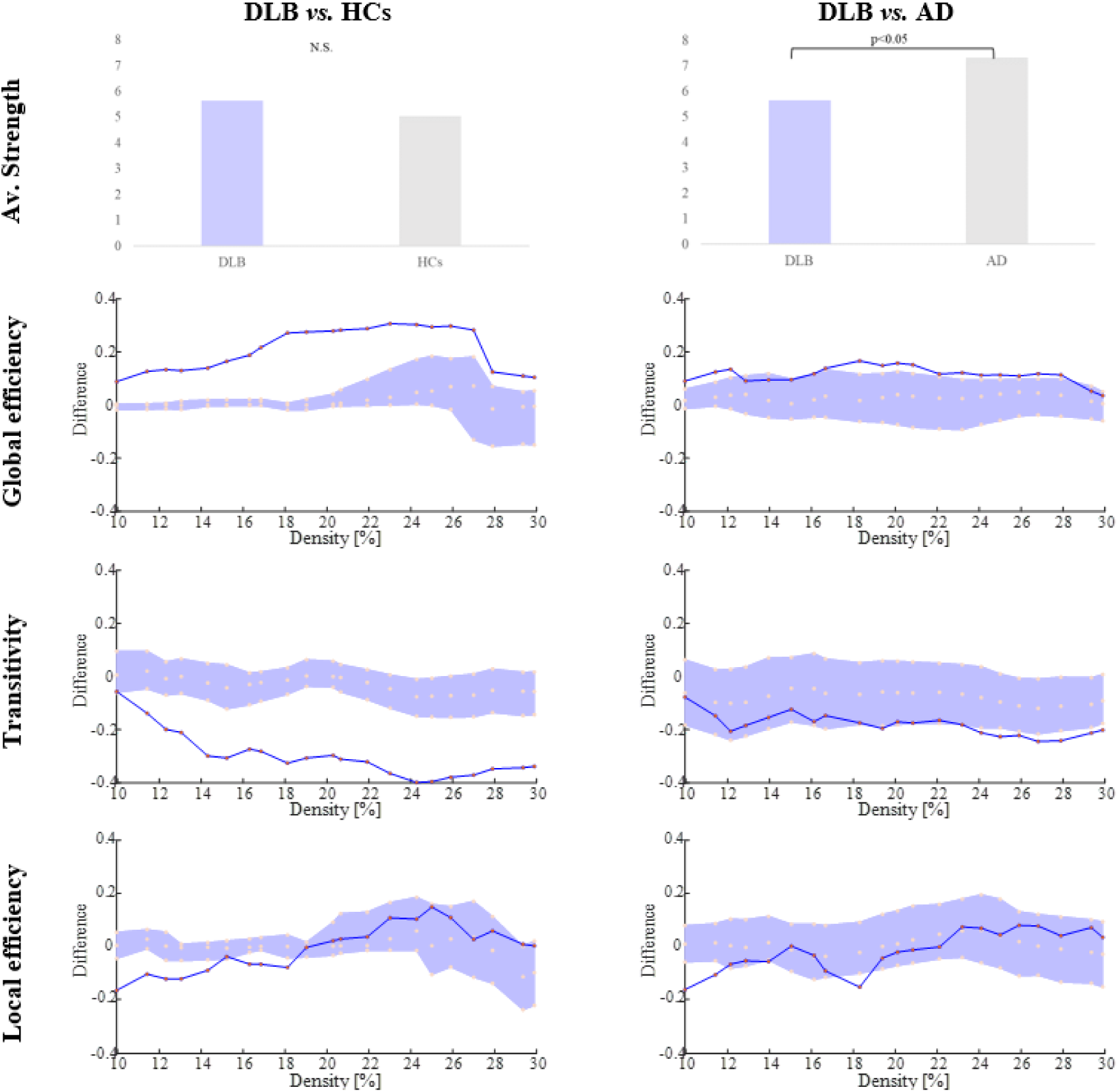
Differences in DLB compared to HCs and AD in global connectome measures. Between-group differences in global connectome measures are displayed on the y-axis. Connectome densities are displayed on the x-axis, spanning from min=10% to max=30%. Red circles refer to the DLB group. Between-group differences are significant when the red circles fall out of the purple-shaded area in 5 consecutive connectome densities. N.S.: non-significant.

### Nodal connectome measures analysis

Table 3 shows the differences in nodal measures across the groups. DLB patients showed a higher nodal strength in an executive variable and a lower nodal strength in a verbal recognition episodic memory variable compared to HCs. DLB patients also had higher participation in an executive variable and lower participation coefficients in visual and verbal memory recognition variables compared with the HC group. Additionally, DLB patients showed higher participation coefficients in another visual and verbal immediate and delayed recall memory variable, and DLB showed a higher nodal integration with higher global efficiency in all cognitive functions, except for one verbal recognition memory variable (and one executive variable (intrusions in phonemic fluency variable) compared with HC. Furthermore, DLB showed a predominantly lower nodal segregation with lower local efficiency in other executive, verbal memory, attention, visuoconstruction, and orientation variables compared with HC. DLB only showed a higher local efficiency in some executive, processing speed, and language variables.

**Table 3.**
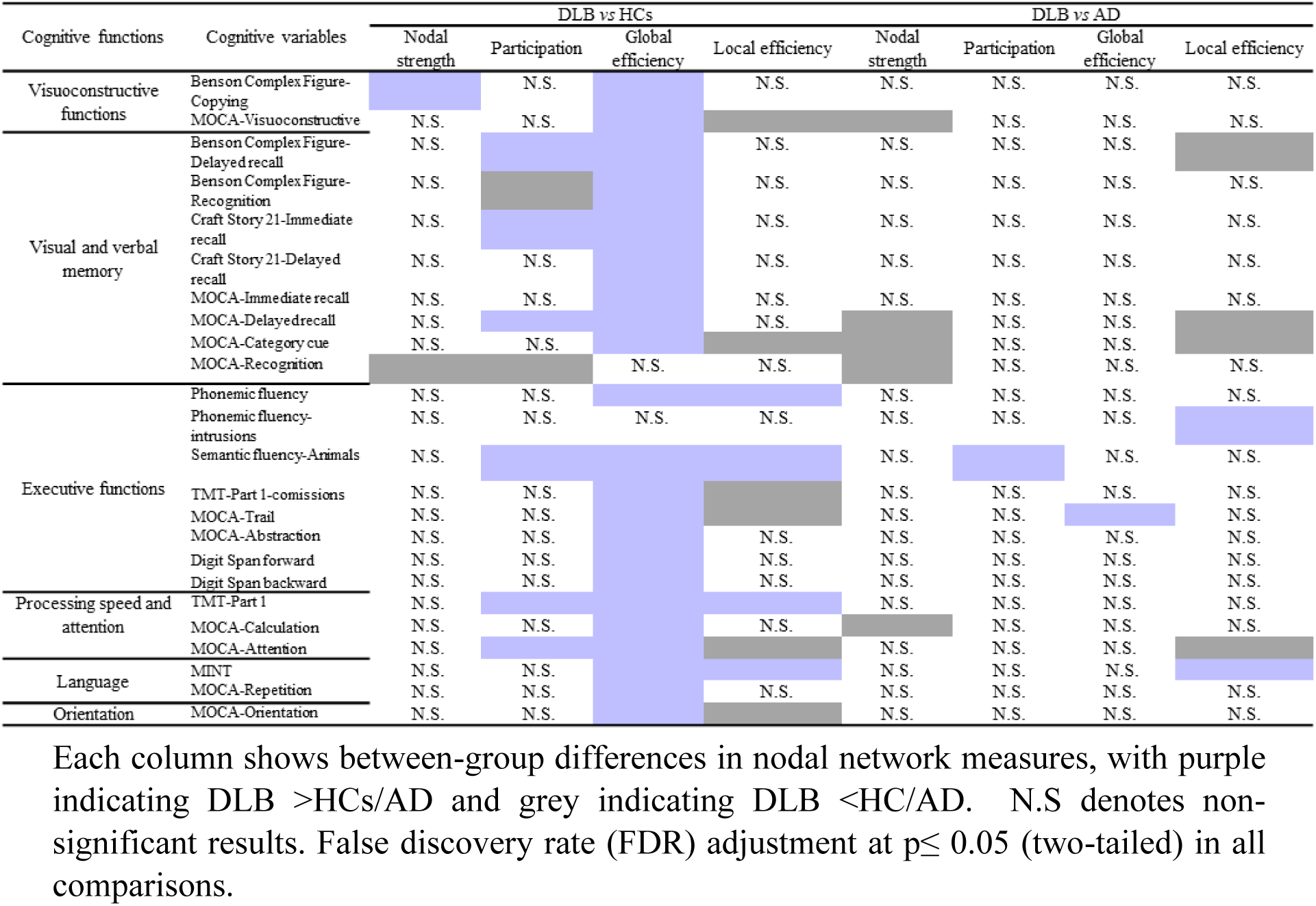
Summary of the differences in nodal measures between groups.

We observed fewer differences in nodal measures when comparing DLB and AD. The DLB group showed a lower nodal strength in several verbal memory variables, a visuoconstructive variable, and an attention variable compared with the AD group. Furthermore, the DLB group showed a higher participation coefficient in an executive variable when compared with the AD group. Moreover, DLB showed a higher global efficiency in one executive variable than the AD group. Additionally, the DLB group showed a higher local efficiency in one language and one executive variable and a lower local efficiency in several verbal and visual memory variables as well as in one attention variable compared to AD.

## DISCUSSION

We characterized the cognitive connectome in DLB patients compared to by HCs and AD patients. Our results showed differences in centrality, integration, and segregation measures for DLB compared to HCs and AD groups.

Our first objective was to characterize the cognitive connectome of DLB patients compared to a HC group. Visual inspection of the cognitive connectome of DLB patients showed that, compared to HCs, DLB patients had a diffuse pattern of uniformly weak correlations within and between cognitive functions. This diffuse pattern was corroborated by quantitative analyses of global and nodal connectome measures, characterized as a less segregated connectome in the context of a higher global efficiency. Because DLB patients performed worse than HCs in all cognitive domains, this combination of segregation and integration (efficiency) findings should be interpreted as a pattern of dedifferentiation (38). The dedifferentiation refers to a higher intercorrelation between cognitive functions and has been associated with reduced neural specificity to cognitive processes (38–40). Previous studies have found that higher global efficiency is related to lower cognitive performance in middle-age and older adults (14,41). The presence of uniformly weak correlations was also evidenced in quantitative analyses at a global and nodal level by the scarce differences found in the centrality measure of strength, which is based on the magnitude of correlations. The diffuse and dedifferentiation pattern found in DLB reflects greater connectome-wide interconnectivity and lower local interconnectivity between neighboring cognitive functions, making it difficult to differentiate segregated groups of cognitive functions in the cognitive connectome of DLB. This dedifferentiation pattern was associated with a worse performance across all cognitive functions in the DLB group. Specifically, the higher integration found in DLB cognitive connectome might reflect a tendency among pathological groups to show concordance in their dysfunction across a range of different cognitive functions (19). Interestingly, two previous functional and metabolic imaging studies also found a dedifferentiation pattern, supporting our findings (42,43) and suggesting that the results of our study could be a reflection of the impairments observed in brain network functioning. However, there are contrary results in other functional or metabolic neuroimaging studies applying graph theory (e.g.45–47). This is a reminder that although some studies have found associations between changes in brain network functioning and cognition (42), these are two different modalities for which one could not expect complete agreement in the results. In this context, complementing results across distinct modalities would improve our understanding of the relationship between brain networks and cognition connectome functioning.

Furthermore, nodal measures pinpointed connectome impairments in specific cognitive functions. Specifically, we observed a higher participation in executive, processing speed, and attention in DLB, which indicates that these cognitive functions have a more prominent role as connector hubs in the cognitive connectome of DLB. Our results are in line with the prominent role of executive functions in regulating other cognitive functions (47), as well as the role of processing speed and attention as central functions that influence other cognitive functions (18,48,49), and expand these results suggesting that impairment in these cognitive functions could be driving the performance in other cognitive functions to a greater extend in DLB compared to HCs. Additionally, the central position of hub nodes in the connectome carries the risk of becoming highly disruptive to the whole connectome in case of failure (50). The fact that attention emerged as an important connector hub in DLB supports the idea that the cognitive fluctuations and the pronounced variation in attention present in the DLB group could have affected the characteristics of its cognitive connectome. The relevance of attention as connector hub highlights the potential of the cognitive connectome to capture the effect of cognitive fluctuations on other cognitive functions in DLB. Moreover, our results on cognitive hubs are in line with some neuroimaging studies showing consistent disruptions in attention networks and their connections to other brain regions (51). Furthermore, we also observed impairments in participation in visual and verbal memory variables in DLB compared to HC. In particular, the memory variables with higher participation coefficients had an important executive component (i.e., retrieval), whereas those with lower participation coefficients were related to consolidation processes. These results are in line with a review that suggested that deficits in recall in DLB are secondary to impairment of executive functions (13) and extend this information, suggesting that memory variables with an executive component could be more central in the cognitive connectome of DLB patients and thus, can be driving or influencing the performance in other cognitive variables to a greater extent than in the cognitive connectome of HCs.

Our second objective was to investigate the cognitive connectome of DLB compared to an AD group. While differences were less extensive than in comparison with HC in the visual inspection of the cognitive connectomes, the global comparisons between DLB and AD support the idea that the pattern of dedifferentiation not only characterizes DLB compared to cognitively normal people but also the AD group. Although previous studies indicated that AD also presents alterations in integration and segregation measures in the cognitive connectome (16,19), our results showed that this alteration seems more pronounced in DLB. This finding suggests that a pronounced dedifferentiation pattern could indicate DLB when compared to AD patients, which is especially relevant as DLB is often misdiagnosed as AD (52). It should be noted that, contrary to findings in the comparison between DLB and HC, we found differences in centrality at a global level between DLB and AD. Specifically, we observed a lower average strength in DLB compared to AD, which may be driven by the presence of generally weaker correlations in DLB, especially in visual and verbal memory functions. This result reinforces the idea that DLB shows a greater pattern of dedifferentiation. Differences in nodal measures between DLB and AD groups complemented the global results. When comparing DLB with AD, we found an alteration in centrality measures at a nodal level, particularly regarding nodal strength. More precisely, we observed a reduced nodal strength in DLB, especially in variables related to consolidation processes in verbal memory (e.g., MoCA-Recognition/Category cue). This reflects the less central role of consolidation processes in DLB as opposed to AD, with the higher impairment in consolidation processes in AD having a greater disruptive effect on the performance in other cognitive functions (8). In this line, previous studies with AD patients and the cognitive connectome analysis approach have reported an important role of memory functions in the connectome of AD and its reorganization in different stages of the disease (16,17,19). However, to our knowledge, this is the first study showing that this is a differential characteristic of the cognitive connectome between DLB and AD. In contrast, we observed less significant differences in centrality when considering the participation coefficient compared to nodal strength. This might suggest that graph measures computed from weighted networks (before binarization, e.g. nodal strength) may be more sensitive to subtle differences such as those between DLB and AD than graph measures computed from binary networks (e.g. participation). Specifically, we observed a higher participation coefficient in one executive variable, suggesting that this aspect is not only a feature of DLB compared with HC but is also differentiating DLB patients from AD patients. Specifically, a semantic fluency variable showed higher participation in DLB compared with both HC and AD. It is worth noting that this variable not only has an important executive component but also performance in this task is determined by processing speed and attention which are typically impaired in DLB. Higher participation coefficients in this variable highlight the prominent role of executive functions as hubs that regulate other cognitive functions, especially in DLB. The significant differences in centrality measures between DLB and AD in our study, underscores the potential of cognitive connectome analysis to characterize the differential cognitive profile between distinct diagnostic groups as well as the relevance of the shift in centrality between cognitive domains to identify cognitive profiles indicative of underlying diseases (19). Furthermore, we found higher global efficiency in the DLB group in one executive variable at a nodal level which, following previous studies, could be related to lower performance (14,41). However, the results in global efficiency in this executive function contrast with the lack of differences observed in the ANCOVA, suggesting that graph theory can capture differences that were not captured by more commonly employed methods in cognition such as univariate analyses. Although we did not find differences between DLB and AD in local efficiency at a global level, our results suggest that there are differences at a nodal level. Of note, we found a predominant reduction of local efficiency in DLB in verbal and visual episodic memory variables. Similar to global efficiency, this reduction in local efficiency may reflect the characteristic higher performance in memory functions in DLB compared with AD (4).

Our findings could have clinical implications. Knowing how cognitive functions are associated with each other and which are central to the cognitive connectome may help improve the challenging differential diagnosis between DLB and AD and help advance in designing more efficient interventions in DLB. Interestingly, a recent systematic review suggested that connectivity measures can potentially become suitable biomarkers for DLB (51). Our results extend this idea and suggest that cognitive connectome measures may also be promising indicators for the presence of DLB, especially considering that cognitive deficits can appear early in the disease (53).

This study has some limitations. The different number of cognitive variables representing each cognitive function may have led to an underrepresentation of some cognitive domains. This study also lacks pathological confirmation for the DLB diagnosis. However, the longitudinal aspect of the NACC database will allow to have pathological confirmation when participants undergo autopsy.

In conclusion, we identified a cognitive connectome in DLB characterized by a dedifferentiation pattern compared with HCs and AD patients. Executive, processing speed, and attention functions played a central role in the cognitive connectome of DLB compared HCs. Furthermore, our data suggest that executive functions and memory functions and their complex associations with other cognitive functions play a distinct role in the cognitive connectome of DLB compared with AD. A novelty of our study is the use of graph theory to investigate the cognitive connectome in DLB. Our findings on the complex organization of cognition in DLB complement alterations in the brain network of DLB patients were revealed by neuroimaging studies. Moreover, our results reinforce the idea that connectivity measures, not only functional MRI but also cognitive, can potentially become suitable DLB indicators in the future. One of the next challenges would be the integration of the cognitive connectome with the more studied brain connectome from structural and functional neuroimaging studies in DLB. Such integration could have relevant implications for research and the clinical field in DLB.

## Data Availability

Data used in this study are available at NACC webpage under request: https://naccdata.org/

## ACKNOWLEDGMENTS

Data used in this study were obtained from the National Alzheimer’s Coordinating Center (NACC) database. The NACC database is funded by NIA/NIH Grant U24 AG072122. NACC data are contributed by the NIA-funded ADRCs: P30 AG062429 (PI James Brewer, MD, PhD), P30 AG066468 (PI Oscar Lopez, MD), P30 AG062421 (PI Bradley Hyman, MD, PhD), P30 AG066509 (PI Thomas Grabowski, MD), P30 AG066514 (PI Mary Sano, PhD), P30 AG066530 (PI Helena Chui, MD), P30 AG066507 (PI Marilyn Albert, PhD), P30 AG066444 (PI John Morris, MD), P30 AG066518 (PI Jeffrey Kaye, MD), P30 AG066512 (PI Thomas Wisniewski, MD), P30 AG066462 (PI Scott Small, MD), P30 AG072979 (PI David Wolk, MD), P30 AG072972 (PI Charles DeCarli, MD), P30 AG072976 (PI Andrew Saykin, PsyD), P30 AG072975 (PI David Bennett, MD), P30 AG072978 (PI Neil Kowall, MD), P30 AG072977 (PI Robert Vassar, PhD), P30 AG066519 (PI Frank LaFerla, PhD), P30 AG062677 (PI Ronald Petersen, MD, PhD), P30 AG079280 (PI Eric Reiman, MD), P30 AG062422 (PI Gil Rabinovici, MD), P30 AG066511 (PI Allan Levey, MD, PhD), P30 AG072946 (PI Linda Van Eldik, PhD), P30 AG062715 (PI Sanjay Asthana, MD, FRCP), P30 AG072973 (PI Russell Swerdlow, MD), P30 AG066506 (PI Todd Golde, MD, PhD), P30 AG066508 (PI Stephen Strittmatter, MD, PhD), P30 AG066515 (PI Victor Henderson, MD, MS), P30 AG072947 (PI Suzanne Craft, PhD), P30 AG072931 (PI Henry Paulson, MD, PhD), P30 AG066546 (PI Sudha Seshadri, MD), P20 AG068024 (PI Erik Roberson, MD, PhD), P20 AG068053 (PI Justin Miller, PhD), P20 AG068077 (PI Gary Rosenberg, MD), P20 AG068082 (PI Angela Jefferson, PhD), P30 AG072958 (PI Heather Whitson, MD), P30 AG072959 (PI James Leverenz, MD). JBT is the Ann and Billy Harrison Centennial Chair in Alzheimer’s Research.

## CONFLICT OF INTEREST

All the authors declared no conflicts of interest relevant to the current study.

## FUNDING

This study was funded by the Swedish Research Council (Vetenskapsrådet, grant 2022-00916), the Center for Innovative Medicina (CIMED, grants 20200505 and FoUI-988826), the regional agreement on medical training and clinical research of Stockholm Region (ALF Medicine, grants FoUI-962240 and FoUI-987534), the Swedish Brain Foundation (Hjärnfonden FO2023-0261, FO2022-0175, FO2021-0131), the Swedish Alzheimer Foundation (Alzheimerfonden AF-968032, AF-980580), the Swedish Dementia Foundation (Demensfonden), the Gamla Tjänarinnor Foundation, the Gun och Bertil Stohnes Foundation, Funding for Research from Karolinska Institutet, Neurofonden, the Foundation for Geriatric Diseases at Karolinska Institutet, and a research fellowship from Government of Canary Islands (call year 2021; TESIS2021010091) co-financed by the Canary Islands Regional Ministry of Economy, Industry, Trade and Knowledge’s Research, Innovation and Information Society Agency and by the European Social Fund (ESF) Integrated Operational Programme of the Canary Islands 2014-2020, Axis 3 Priority Theme 74 (85%).

## DATA AVAILAVILITY

Data used in this study are available at NACC webpage under request: https://naccdata.org/

## Notes

### Competing Interest Statement

The authors have declared no competing interest.

### Author Declarations

Institution Review Board of Alzheimer's Disease Research Centers included in the National Alzheimer's Coordinating Center database gave ethical approval for scientific works derived from the use of data in the National Alzheimer's Coordinating Center database.

