## Supplementary figures and table for "The Cognitive Connectome in Dementia with Lewy Bodies"

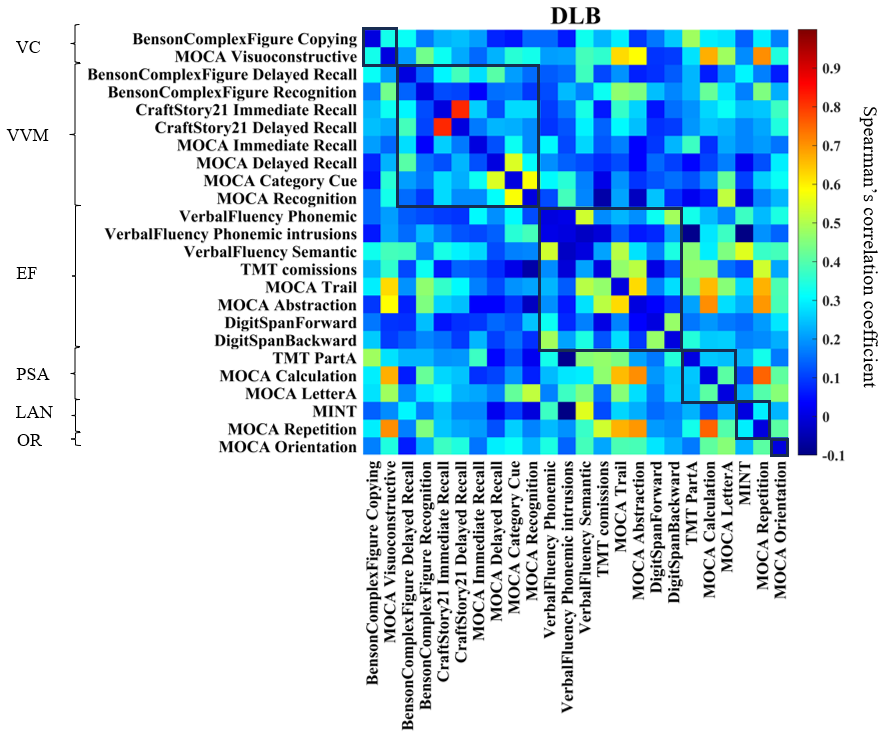

**Supplementary figure 1. Correlation matrix of DLB group**. Cognitive connectome of DLB grouped by cognitive functions. TMT, Trail Making Test; MoCA, Montreal Cognitive Assessment; MINT, Multilingual Naming Test. VC, visuoconstructive functions; VVM, visual and verbal episodic memory; EF, executive functions; PSA, processing speed and attention; LAN, language; OR, orientation.

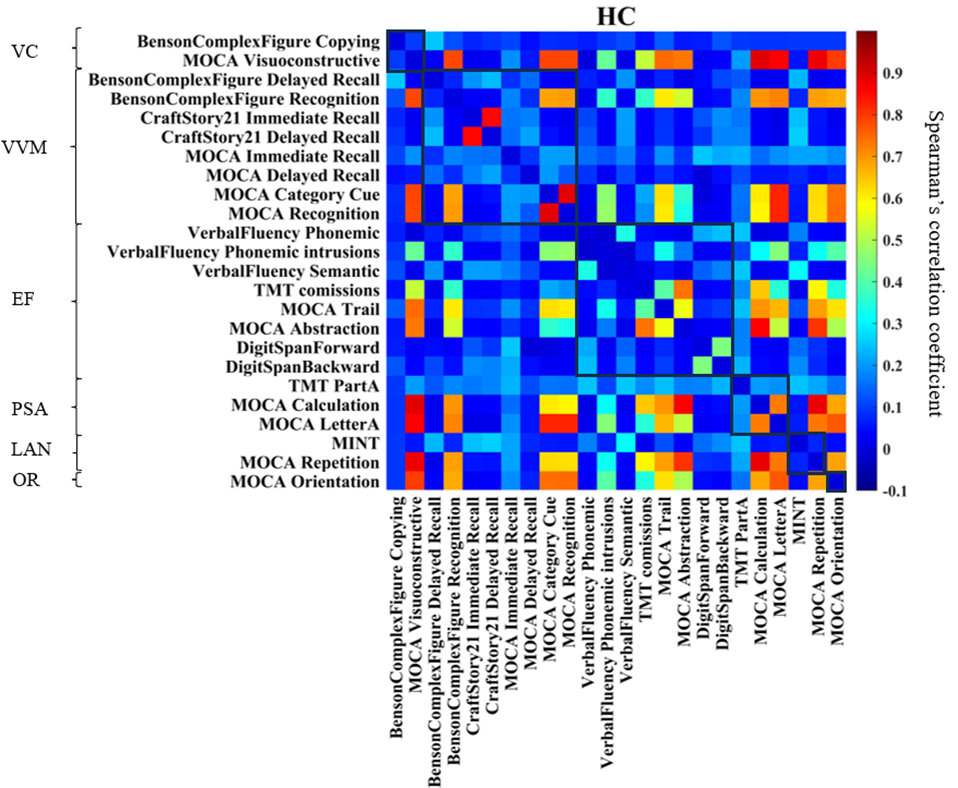

**Supplementary figure 2. Correlation matrix of HC group.** Cognitive connectome of HC grouped by cognitive functions. This matrix did not show negative correlation, but the range of color bar numbers was equalized across all groups to facilitate comparison. TMT, Trail Making Test; MoCA, Montreal Cognitive Assessment; MINT, Multilingual Naming Test. VC, visuoconstructive functions; VVM, visual and verbal episodic memory; EF, executive functions; PSA, processing speed and attention; LAN, language; OR, orientation.

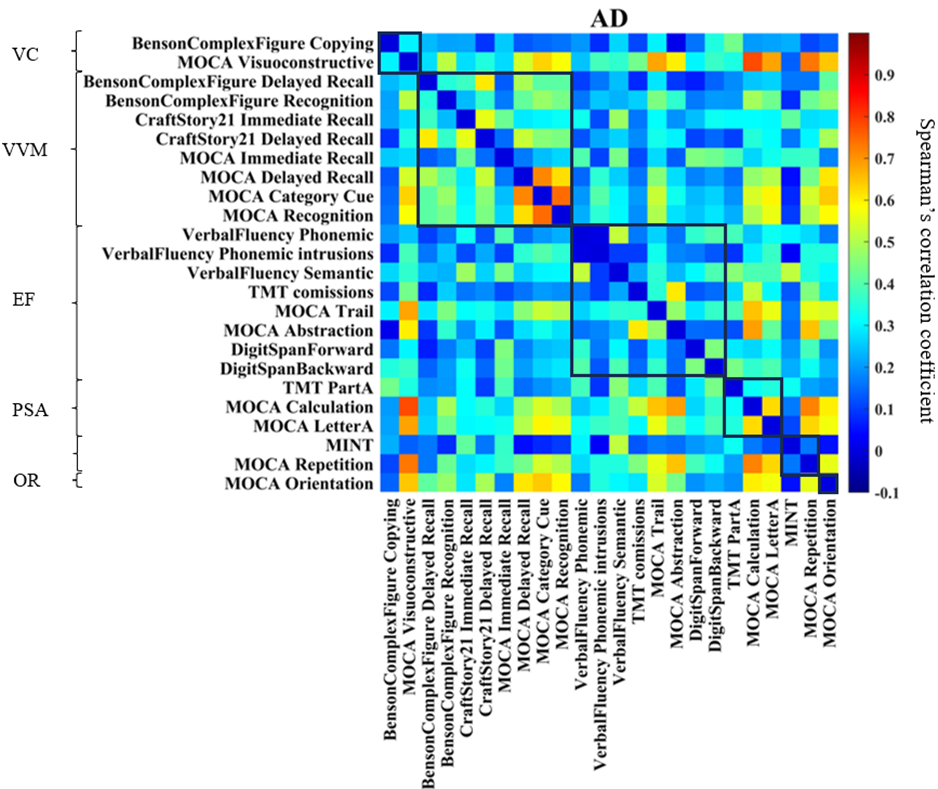

**Supplementary figure 3. Correlation matrix of AD group.** Cognitive connectome of AD grouped by cognitive functions. TMT, Trail Making Test; MoCA, Montreal Cognitive Assessment; MINT, Multilingual Naming Test. VC, visuoconstructive functions; VVM, visual and verbal episodic memory; EF, executive functions; PSA, processing speed and attention; LAN, language; OR, orientation.

**Supplementary Table 1. Graph theory measures**

| Measures | | Definition and relevant comments |
| --- | --- | --- |
| Global efficiency | Nodal global efficiency (54) | Average of the inverse shortest path length between a specific node and the rest of the network. |
|  | Average global efficiency (54) | Average of the global efficiencies of all nodes. It measures how efficiently information is exchanged throughout the network. This measure, in contrast to de characteristic path length, can be computed on disconnected networks (55) |
| Local efficiency | Nodal local efficiency (54) | Global efficiency of a node calculated on the subgraph created by the node’s neighbors. |
|  | Average local efficiency (54) | Average of the local efficiencies of all nodes. |
| Strength | Nodal strength (56) | Sum of the weights of all edges connected to a node. |
|  | Average strength (34) | Average of the strengths of all nodes. |
| Transitivity (34) | | Fraction of a node’s neighbors that are also neighbors of each other in the whole network, normalized by the whole network, reflecting how well the nodes are connected to nearby regions forming cliques. The transitivity measure is similar to the clustering coefficient but is less vulnerable to methodological issues (e.g., edge definition, network size, and groups composition) (33,57) |
| Participation (37) | | Quantifies the relation between the number of edges connecting a node outside its community and its total number of edges. |
